# Evaluation of the implementation and effects of management through care and services pathways: A protocol study

**DOI:** 10.1101/2023.05.25.23290549

**Authors:** Lara Maillet, Georges Charles Thiebaut, Anna Goudet, Isabelle Godbout, Nassera Touati, Pernelle Smits, Arnaud Duhoux, Mylaine Breton, Sabina Abou-Malham, Yves Couturier, Frédéric Gilbert, Jean-Sébastien Marchand, Jean-Louis Denis

**Affiliations:** École Nationale d’Administration Publique (ENAP) Montréal (Québec), Canada; Institut universitaire de première ligne en santé et services sociaux (IUPLSSS), CIUSSS Estrie-CHUS, Sherbrooke (Québec), Canada; Faculty of Business Administration, Université Laval, Québec (Québec), Canada; Faculty of Nursing, université de Montréal, Montréal (Québec), Canada; Faculty of Medicine and Health Sciences, Université de Sherbrooke, Sherbrooke (Québec), Canada; Faculty of Medicine and Health Sciences, School of nursing, Université de Sherbrooke Sherbrooke (Québec), Canada; School of Social Work, Université de Sherbrooke Sherbrooke (Québec), Canada; School of Management, Université du Québec à Montréal Montréal (Québec), Canada; Department of Health Policy Management and Evaluation, School of Health, Université de Montréal, Montréal (Québec), Canada; Commissaire à la santé et au bien-être (CSBE) Montréal (Québec), Canada

## Abstract

**Background:** In 2015, the Government of Quebec undertook a vast reorganization of its health and social services network. This reform mainly aimed to promote and simplify access to services for the population, contributing to the improvement of the quality and safety of care, and increasing the efficiency and effectiveness of the network. Since 2016, several health care organizations (HCOs) have pushed reform even further by developing management through care and service pathways (MCSP). This study aims to identify, in a processual manner, the different factors involved in implementing MCSP in different HCOs, in the turbulent context of the COVID-19 pandemic.

**Method:** The methodology of this research project is based on developmental evaluation. The objective of developmental evaluation is to guide organizations and actors in the adaptation and development of innovations in complex and turbulent environments. Data will be collected over a three-year period using five strategies: i) organizational questionnaires; ii) analysis of clinical-administrative databases; iii) documentary analysis (grey and scientific literatures); iv) participant observations and v) semi-structured interviews with key actors involved in the implementation of MCSP.

**Discussion:** In addition to the operationalization of pathways, the implementation of MCSP i) involves transforming the governance of the health care organization both at the strategic and operational levels and ii) is a demanding process that requires changes in practices, modifications in the allocation and configuration of resources and the development of new collaborations between the different actors in the organization, the partners and the users involved in this transformation. Several studies claim that governance innovations can create conditions that are favourable to the emergence of innovations in terms of available services and responding to the needs of populations. This research will develop knowledge of the factors involved in implementing MCSP in complex and turbulent contexts and propose scale-up across the province.

## Introduction

### Background

Ensuring the enhanced organization of services in order to promote access appears to be a priority for many health care systems, both in Canada and at the international level (1, 2). To that end, many have developed methods of management through care and service pathways (MCSP) (3, 4), which address the complexity of the issues faced by health care systems: accessibility, equity, performance, and allocation of resources, among others. Indeed, MCSP represents a form of governance innovation that aims to increase the capacities of health and social services organizations to adapt to the needs of the population, notably through the improvement of accessibility and the continuity of services (5). MCSP therefore aims to improve the performance of HCOs on an ongoing basis, through greater integration of services and practices, by following four guiding principles: ongoing improvement, partnerships, measurement of performance by choosing appropriate monitoring indicators, and the establishment of a matrix governance model based on a bottom-up approach (6).

This research project is based on a pilot study carried out in 2017-2019 (6) and aims to monitor the development and implementation of the MCSP to enrich the body of knowledge and feed the teams in charge of this implementation. However, several researchers raise the difficulties to implement pathways which represents a major organizational change, requiring significant technological and administrative means and resources (7). In addition to the operationalization of pathways, 1) implementation of MCSP involves transforming the governance of the HCO both at the strategic and operational levels, and 2) is a demanding process that requires changes in practices; modifications in the allocation and configuration of resources; and the development of new collaborations between the different actors in the organization, the partners and the users involved in this transformation. This is why considering the contexts for implementation and allowing for the adaptation of processes are some of the challenges revealed by past pandemic experiences (8, 9) and the current COVID-19 pandemic, which poses a major and unprecedented challenge to health care system (10).

In this regard, there are four major gaps between the field of MCSP and the scientific literature in this domain. First, enhanced understanding is needed of how MCSP, as a new method of matrix management, influences HCOs both in terms of clinical and managerial practices and governance. Second, what are the facilitators and barriers to the implementation of MCSP, both at the micro level (organization and territory) and at the macro level (scaling to other contexts)? Third, what are the most appropriate indicators to take MCSP into account at the clinical, managerial, and financial levels, as well as in terms of equity and co-evolutionary links with partners that are external to the organization? Fourth, there is a real need to better theorize the implementation of MCSP, considering the interdependence with the context (partners external to the organization) and the role of adaptation in the large-scale implementation of an innovation such as MCSP.

#### What do we mean by management through care and service pathways in health care organizations?

Several definitions and names are used in relation to pathways: care pathways; care process effectiveness; clinical pathways; critical pathways; integrated care pathways; case management plans; clinical care pathways; care maps (11). Notably, we can refer to the definition adopted by the European Pathway Association (EPA):

> “A complex intervention for the mutual decision making and organization of predictable care for a well-defined group of patients during a well-defined period. Defining characteristics of pathways include: an explicit statement of the goals and the key elements of care based on evidence, best practice and patient expectations; the facilitations of the communication and coordination roles and sequencing the activities of the multidisciplinary care team, patients and their relatives, the documentation, monitoring and evaluation of variances and outcomes; and the identification of relevant resources.” p.3, (12).

The scientific literature indicates that the pathways can take different forms, from the description of actual practices and tools to improve them (e.g. a document that describes or supports the coordination of services throughout the care process) to more prescriptive strategies (e.g. a care plan, a procedure/process, a guide to best practices) that are very detailed, even complex (11-13). They can cover different themes (e.g. a population suffering from a pathology, or an action/process), but must be based on clearly identified inclusion/exclusion criteria (14). For example, the following inclusion criteria are often cited: reach a significant number of users; target the critical mass (60 to 80% of a targeted group); choose a pathway with a high level of predictability that requires a certain level of consistency in its practices; choose a pathway with a high cost of care; prioritize a pathway requiring multidisciplinary care (14-16).

Certain pathways can be based on the actual situation and developed through a continuous improvement loop, or on a revised version considered as a best practice. All require the establishment of a clinical and organizational integration process at all levels of the organization (14) allowing support of the ongoing improvement, quality and efficiency of care (11). Based on multidisciplinary work both in terms of its design and operations (9, 13), the pathways could be described as:

- Supporting the formalization of the sequencing of care processes and actions and, in some cases, their standardization (13, 16);
- Supporting shared decision making (9);
- Reducing variations in care, both in terms of the route towards accessing care and in practices (or interventions);
- Increasing the quality of care in terms of relevance, competent implementation, and security (9, 11, 13).

With regard to coordination, Vanhaecht, De Witte and Sermeus (2007) propose three models which they position based on lines of predictability as well as consensus between the members of a multidisciplinary team (12). These models are:

- the Chain model: adapted to high levels of predictability and consensus, where there is a very high degree of precision for each stage and the time between the stages is relatively short (e.g. planned surgery, chemotherapy treatment);
- the Hub model: structured around a case manager for a moderately high level of predictability and consensus, where flexibility or a long monitoring period is necessary in certain cases (e.g. internal medicine, rehabilitation, psychiatry);
- the Web model: adapted to a high level of unpredictability, where the regular (i.e. daily) adjustment of a professional network linked together by an information system and coordination mechanisms is necessary (e.g. crises, emergencies, intensive care of users with multiple comorbidities) (13).

### Study context

In 2015 the Government of Quebec undertook extensive reorganization of its health and social services network by establishing merged large health care organizations (HCOs) with more than 10,000 employees that bring together several HCOs with different missions on several sites and covering large geographical territories. Since 2016, several of these large HCO have pushed reform even further through development of MCSP. The latter is a *“operating method based on structural foundations and criteria allowing the optimization of care and services pathways to ensure they adhere to the needs of users”* p.13, (17). In turn, care and services pathways are *“the part of the process followed (care interventions and episodes) by a group of users with a clinical condition or a similar profile […], they integrate, in a cross-cutting manner, the access mechanisms, interventions for the promotion of health, evaluations/research/guidance, follow-up/support treatment by way of preventative treatment”* p.15, (17).

Since 2017, the research team has been following the implementation of the MCSP in one and then three HCOs that have developed the MCSP in their own way, depending on their geographic, socioeconomic, populational and epidemiological context since the reform in 2015. In 2018, the Ministry of Health and Social Services (MSSS) mobilized these 3 HCOs to form a provincial committee to conceptualize and implement an integrated model of MCSP fed by the experiences of each of these establishments. This new ministerial model has been implemented in these three “developer” HCOs as well as in three other establishments “experimenter”. These three HCOs are identified as “experimenter” because they did not participate in the initial implementation process. Since, the research team has documented and evaluated the implementation of MCSP in Quebec, initially in these three “developer” HCOs. This research project aims to continue the evaluation of the three “developer” HCOs as well as the scaling up of the three “experimenter” HCOs. The main lessons learned from the pilot study show that in Quebec, MCSP innovation is characterized by two main elements and two main consequences (6).

Firstly, an activity phase based on the strategic communities approach (18, 19). Strategic communities are interorganizational collaborative structures composed of professionals, frontline managers, general practitioners, medical specialists and representatives of community organizations (whose mandate is to create, implement and evaluate new ideas relating to the organization of intersectoral work (18, 19). This means that MCSP is based on the collaboration and cooperation of all the actors involved in health care and service pathways and in which both users and community partners (e.g., community organizations, municipalities, schools) are closely involved. With this perspective, the operating methods are no longer hierarchical, but rather cooperative, with a significant part left to the emergence and joint creation of diagnoses and solutions (10). Involving users and community partners from a systemic perspective is part of this theoretical reflection surrounding the MCSP.

Secondly, this innovation needs to introduce a matrix structure into the organization to link the facilitation structure and the management structure. The individuals in charge of MCSP are functional specialists who are responsible for the implementation, functioning and improvement of care and services pathways through the creation of new organizational methods for services (20). Horizontal coordination across the matrix (the pathways) and between the horizontal and vertical lines (the directions) is done through mutual adjustment between the administrative and clinical professionals at all levels of governance (20). This type of structure aims to profile the service offer and increase the flexibility and cross-cutting nature of the organization to adapt it in a way that is relevant to contextual needs (of the users, the partners, the population of the territory, etc.). Thirdly, MCSP involved the change of vision from traditional management to a large-scale matrix management model, while respecting the specific context of each organization, which was a significant challenge to overcome.

Finally, implementing MCSP means the development of new roles. Dual collaboration and the sharing of power and leadership (between health professionals, including doctors) are some of the additional challenges faced.

These findings lead us to define MCSP as a governance innovation based on a cross-sectional, non-hierarchical approach to the operation of health organizations, where collaboration with external partners and users is central to ensuring the integration of services aimed at improving the health of the population in a given territory (6).

### Conceptual framework

The implementation of MCSP will be analyzed as a governance innovation whose objective is to improve the integration of health care and services with the involvement of partners in the territory, with a view to improving access for the entire population being served to improve access to the entire population served. Such implementation requires a systemic and territorial vision. The proposed framework constitutes a theoretical contribution in the field of implementation. In addition to integrating the five characteristics influencing the implementation of an innovation, it incorporates the notion of multilevel governance as well as a model for assessing the effects of the pandemic on the governance of MCSP. Although a conceptual model such as the Consolidated Framework for Implementation Research (CFIR) (21) could have been used, the present theoretical construct better fits the context of the study and the governance adopted by the MSPC in HCOs. Nonetheless, the CFIR was used in the construction of the Performance Analysis Model on which we rely for the development of our questionnaires.

Fig 1 shows the interlinking of these innovation characteristics and the interrelated dimensions for integration through MCSP implementation process in each context. The pandemic context is also one of the elements considered. This framework that builds on previous studies (21), is composed by four theoretical main elements:

1. Based on the Complex Adaptive System (CAS), which involves a high number of actors or components that are different, autonomous, and above all interdependent (22-24). Their interdependence is what interlinks the influence of an actor or a component with the presence and the intensity of the presence of other actors or components. From this CAS perspective, the use of multilevel governance can be encouraged through the combination of operational autonomy and interdependence between the organizations and systems (21). This multilevel governance is envisaged in a pluralist context, in which it is impossible to conceive of an absolute authority because of shared leadership and distributed powers (25, 26). This leads us to think of health care organizations in terms of interactions and co-evolution between the actors, going beyond a vision that is merely hierarchical (27).
2. There are five characteristics influencing the implementation of innovation as defined by Greenhalgh et al. (2004) (28) and Klein et al. (1996, 2001) (29, 30): (i) the characteristics of the actors (perceptions, values); (ii) the attributes of the innovation (ease of use, correspondence with practical needs); (iii) the internal characteristics of the organization in which the innovation is being implemented (the availability of resources, the adaptation of structures); (iv) the characteristics of the external environment (implication and effects on the partners, co-evolutionary principles); (v) the characteristics related to the piloting of an innovation (strategy and implementation process). Together, these elements will enable focus on the factors that are key to success in the implementation of pathways and identification of the essential elements to ensure their transferability. In doing so, it is crucial to take the interaction between these different elements into consideration using the CAS framework.
3. The conceptual framework includes the dimensions of the integration process that aim to establish coherence between the clinical system, multilevel governance and the collective system for the interpretation and values that form the space in which the actors interact (31, 32). These dimensions are clinical integration; integration of the clinical team; functional integration; normative integration and systemic integration.
4. The framework for robust governance strategies in the turbulent context of the COVID-19 pandemic. Because of the COVID-19 health crisis, we have decided to add the model for robust governance strategies in a turbulent context developed by Ansell, Sorensen and Torfing (33). This framework will be used to gain a detailed understanding of the effects of COVID-19 on the governance of MCSP in the HCOs being studied, as well as on their network of partners. The implementation of the MCSP is the implementation of a particular modality of process management. The governance innovation in the case of this implementation is the implementation of a matrix governance within the HCOs (34). This matrix governance allows to manage in a transversal and therefore processual way: it is the type of governance allowing a process management such as the MCSP. To analyze the organizations being studied, we will use the typology of the six strategies proposed to formulate robust governance solutions. Robust governance strategies aim to maintain or achieve a public program, function, or purpose through the flexible adaptation, agile modification, and pragmatic redirection of governance solutions. Table 1 presents the strategies proposed for the formulation of robust governance solutions (33). The inclusion of a COVID-19 component as part of our research will allow us to understand *in situ* the robust governance strategies implemented in the current turbulent context of the COVID-19 pandemic, which has had a major impact on public services such as the health care system in terms of adaptation and change (33). The pandemic has caused pathways to be altered, transformed or mature enough to continue “unsupervised” (self-organized). This study will document and analyze these phenomena.
5. In order to meet objectives 2 (conditions for implementing MCSP), 3 (measurement of the impacts of the pandemic on the MCSP as well as the possible effects of the MCSP), 4 (analysis of the performance of the MCSP) and 5 (influence context and partnership networks), the team will rely on the Performance Analysis Model (PAM) co-developed by Thiebaut, Maillet and the provincial committee (42). This analytical framework includes 11 dimensions (see table 2).

**Fig 1:** Conceptual framework.

**Table 1:** Strategies proposed for the formulation of robust governance solutions.

| **Terms** | **Definitions** |
| --- | --- |
| **Scalability** | "Aims to flexibly mobilize and de-mobilize resources across organizations, levels and sectors to scale the provision of particular solutions to meet changing needs and demands (35)." |
| **Prototyping** | "Aims to create new, adaptative solutions through iterative rounds of prototyping, testing, and revision based on prompt feedback (36)." |
| **Modularization** | "Aims to create solutions that are divided into a series of modules that can be used flexibly in response to changes in the different aspects of the problem at hand (37)." |
| **Bounded autonomy** | "Aims to create a broad-based ownership and strategic commitment to an overall strategy by involving regional and local actors in the implementation of key tasks and regulations and encouraging them to adapt the overall governance strategy to the changing conditions on the ground (38)". |
| **Bricolage** | "Aims to flexibly use and combine available ideas, tools, and resources to fashion a workable solution in the face of turbulence (39, 40)." |
| **Strategic polyvalence** | "Aims to deliberately design solutions that can be taken in new directions and serve new purposes depending on situational analyses of demands, barriers, and emerging opportunities (41)." |

## Materials and Methods

### Study aims

This project represents an opportunity to follow and support the implementation of a scale-up of a governance innovation from its point of departure, within four HCOs in real time. More specifically, the project aims to:

**Table 2:** Performance Dimensions and Indicators.

|  | Performance Dimensions | Possible indicators |
| --- | --- | --- |
| **Structure and resources** | **Viability** | Number of nurses per 1,000 inhabitants (or according to the total number of people targeted by the trajectory) [11] for the Territorial and local service networks (RTS) where the trajectory is implemented |
|  | **Accessibility** | Rate of use of services by specific populations (defined according to language, age, origin, etc.) over a given period<br>Number of days before the 3rd available appointment |
|  | **Adjustment to the needs of the target population** | Age-standardized rate of mental health hospitalizations per 100,000 inhabitants<br>Adjusted rate of hospitalizations for ambulatory care sensitive conditions per 100,000 inhabitants under age 75<br>Proportion of people aged 65 and over with loss of autonomy reached through home support services and having benefited from an intervention for the prevention of falls |
| **Process** | **Coordination and continuity** | Percentage of clients followed in oncology by a designated pivot nurse in oncology (PNO), regardless of the place of intervention<br>Rate of emergency hospital admissions of patients due to acute exacerbations of their chronic disease |
|  | **Relevance** | Proportion of family physicians who report that they consistently provide written instructions on how to manage their home care to their patients with chronic conditions [10]<br>Proportion of clinicians using clinical practice guidelines for certain pathologies [10] |
|  | **Security** |  |
|  | **Humanism** | • Time spent with patients in terms of average duration of a regular consultation [11]<br>• Proportion of workers met by a user and who clearly communicated their role |
|  | **Productivity** | • Rate of use of operating rooms<br>• Average length of stay for the eight main causes of high-volume hospitalizations (according to the trajectories chosen and the references for each of these trajectories) |
| **Outcomes** | **Clinical and population efficacy** | CLINICAL:<br>• In-hospital death rate within 30 days of major surgery<br>• Adjusted rate of hospitalizations for ambulatory care sensitive conditions per 10,000 population (e.g., COPD, diabetes)<br>POPULATION:<br>• Rates of regular and occasional smokers by age and gender [27]<br>• Proportion of the population with obesity [27] |
|  | **Efficiency** | • Average cost per screened case of breast cancer [12]<br>• Percentage of public drug program spending on generic drugs |

1. Document the clinical, organizational and partnership practices established by MCSP in each participating HCO (developers and experimenter), based on the four principles of MCSP (ongoing improvement, partnership, measurement of performance, and the establishment of a matrix governance model);
2. Describe and analyze the implementation conditions over time (facilitators and barriers) of the structure and the process of MCSP for each of the HCOs (“developer” and “experimenter” HCOs);
3. Measure the effects of the implementation of MCSP on the degree of integration of care and services for the users, the partners, and the HCOs;
4. Analyze and measure the effects of the COVID-19 pandemic on MCSP and, in turn, how MCSP has been able to contribute in terms of the adaptation of “developer” and “experimenter” HCOs in light of the pandemic;
5. Understand the influence of the context, including the management of the pandemic, on the implementation of MCSP and on its effects.

### Design study

The methodology of this research project is based on developmental evaluation. The objective of developmental evaluation is to generate ongoing knowledge to guide organizations and actors in the adaptation and development of innovations in complex environments (43). The evaluation allows for analysis of the implementation and its effects, conducted through the implementation of projects while taking into consideration the needs of the users of the innovation, at the operational, tactical, and strategic levels as well as at the contextual level (partners). We will conduct a multilevel and multiple case study using process analysis (44-46). This method consists of synthetic research by multiple cases study (30) with nested levels of analysis to describe, measure and analyze the implementation of the MCSP in different contexts (47, 48) As part of our study, we propose a longitudinal study of interlinked cases (49, 50) to understand and to compare intra- and inter-organizational variations at a given moment and over time (50).

### Case study

The three “developer” HCOs were selected in 2018 by the MSSS to participate in the provincial committee and in the development of a scaling-up of MCSP through their own experiences (Table 3).

**Table 3:** Characteristics of the three “developer” HCOs.

| HCO | Geographical region | Population and territory | Number of employees | Number of pathways to follow | Pathways/ continuum followed (tracer cases) |
| --- | --- | --- | --- | --- | --- |
| A | Semi-urban and rural | 476,000<br>(2015) 12,820 km<sup>2</sup> | 17,000 | 2 | Chronic diseases<br>Alternate level of care (ALC) |
| B | Very urban | 445,600<br>(2018) 88 km<sup>2</sup> | 12,369 | 2 | Chronic diseases<br>Healthy weight management |
| C | Rural | 516,281<br>(2018) 47,000 km<sup>2</sup> | 17,211 | 2 | Chronic diseases<br>Autism spectrum disorder (ASD) |

The “experimenter” HCO will be selected by our research team in accordance with their interest in experimenting with MCSP, the level of integration of MCSP already achieved (variability of the initial conditions for implementation) and the diversity of contexts between cases (Table4).

**Table 4:** Characteristics of the “experimenter” HCO.

| HCO | Geographical region | Number of trajectories to follow | Pathways/ continuum followed (tracer cases) |
| --- | --- | --- | --- |
| D | Urban | 2 | Chronic diseases<br>Alternate level of care (ALC) |

In each of the HCOs we will follow two tracer cases, specifically two pathways (care and service continuums), to study the implementation of MCSP (Objectives 1 and 2); that of the Performance Analysis Model for MCSP (Objectives 3 and 4) and the effects of this implementation (Objectives 4 and 5).

### Data collection

We will use five sources of data: documentary analysis, participant observation, semi-structured individual interviews, organizational questionnaires, and analysis of clinical-administrative databases. The data collected as part of this research will not be made available for replication because it is confidential and restricted by ethical considerations.

#### Qualitative data

We will carry out documentary analysis of the available written records on the structure and process of MCSP for each case under study. These documents will come from the gray literature collected during the three years of implementation of the pilot project for the implementation of the MCSP in the three “developer” HCOs and with the provincial committee. In addition, we will conduct participant observations during the meetings that will take place at the different levels. For example, participant observations will be made during meetings of operational visual stations (working group, and pathway coordination body). They will be undertaken by the members of the research team and recorded in the form of field notes, taking descriptive account of the development of the different situations observed. We plan to conduct 44 participant observations (12 per “developer” HCO and 8 per “experimenter” HCO).

We will also carry out individual semi-structured interviews. The interviews will focus on the implementation of MCSPs and their effects on the operation and degree of integration of care and services. These will be undertaken by at least one researcher and one student, in person, by videoconference or by telephone, with the key actors involved in the implementation of MCSP, until data saturation is achieved (see Table 5; N anticipated: 78 to 81). The selection of key participants will be based on the methodology of reasoned and selective choices (51) and will reflect the diversity of the actors involved in the pathway coordination bodies or in decision making, including users, community partners, practitioners, managers, senior executives and ministerial actors. As part of the institutional convenience committee request, a gatekeeper per HCO has been identified to support the research team in carrying out the project. Institutional convenience consists, among other things, in verifying the adequacy between the clientele targeted in the research project and the clientele of the establishment, the availability of the facilities, equipment and human resources of the establishment. The gatekeeper will connect potential participants with the research team. Initial contact and recruitment will be made by the principal investigators or research professionals via email or telephone.

**Table 5:** Type and estimated number of participants.

| Type of participants | Number of interviews (estimated) |
| --- | --- |
| **Users** who are members of pathway coordination bodies | $N = 2 \times 2 \times 4 = 16$ users |
| **Community partners** who are members of pathway coordination bodies | $N = 1 \times 2 \times 4 = 8$ partners |
| **Local practitioners/managers** who are members of pathway coordination bodies and operational visual sites | $N = 2 \times 2 \times 4 = 16$ practitioners |
| **Managers** responsible for tactical and strategic pilot projects | $N = 2 \times 2 \times 4 = 16$ managers |
| **Senior executives** present in the strategic pilot project room | $N = (2 \times 4) + (2 \times 4) = 16$ decision makers |
| **Ministerial actors** connected to MCSP and its implementation | $N = 3$ individuals from the MSSS |
| **Pan-Canadian actors** involved in a similar process | +/- 3 to 6 |
| **Estimated total interviews** | **78 to 81** |

#### Participants and recruitment

According to the governance structure currently envisaged (Fig 3), the following participants were anticipated for the individual interviews (Table 5).

**Figure 2:**
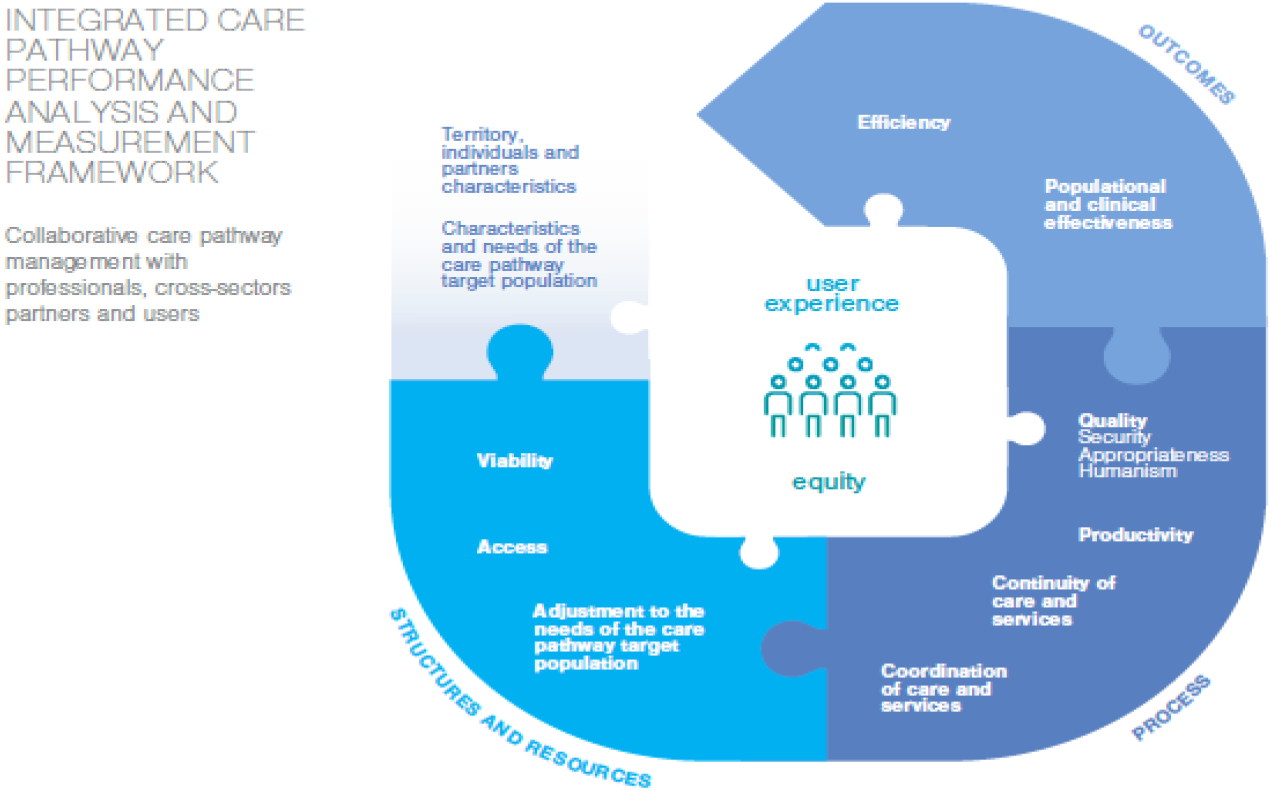
Integrated care pathway performance analysis and measurement framework (41)

**Fig 3:** Generic matrix governance structure for MCSP (from working draft)

#### Survey and database

The quantitative component includes two parts for data collection and analysis: i) an organizational questionnaire, and ii) the consultation of clinical-administrative databases.

We will develop an organizational questionnaire to measure the effects of MCSP on matrix governance processes and operations, as well as the effects of MCSP on accessibility, continuity and equity of the care and services provided by each of the HCOs under study. This survey is based on the Performance Analysis Model (PAM) for MCSP, co-developed by Thiebaut, Maillet and the provincial committee (42) (see fig. 2 and table 2). It will be sent to the four HCOs on four separate occasions between 2023 and 2025. These four rounds of data collection will allow us to gain an organizational picture of MCSP both during and after the pandemic context. A purposeful sampling will be carried out to identify the participants who will answer the questionnaire (Table 6).

**Table 6:** Type and estimated number of participants.

| Type of participants | Number (approximative) |
| --- | --- |
| Organizational survey: managers, senior executive, clinicians, intervenants | $N = 40 \times 4 = 160$ |
| Partners (community) survey | $N = 4 \times 10 = 40$ |
| User survey (healthcare users) | $N = 4 \times 10 = 40$ |
| Total | $N = 240$ |

To analyze the profiles of use by targeted pathways, we will refer to two clinical-administrative databases: iCLSC and InfoCentre in the HCOs that are relevant to this study. Insofar as possible, we will use data that has already been collected and monitored by the pathway management bodies.

The variables requested per pathway will be (non-exhaustive list): age; gender; place of birth; place of residence; diagnosis; type of treatment; type of services accessed; professionals consulted; referencing and other data allowing the establishment of the profile types for each pathway studied as well as the monitoring of the performance indicators of the PAM (accessibility, continuity, and equity). All relevant data collected by the MCSP bodies will also be assembled. Being in a co-construction process (developmental evaluation), we have access to anonymized and aggregated clinic-administrative data as soon as we are involved in the field.

### Data analysis

Analysis of the multiple data sources will require a combination of qualitative and quantitative approaches.

#### Qualitative data analysis

The qualitative data will be collected through a documentary analysis, participant observations and semi-structured individual interviews. The individual interviews will be recorded and transcribed. The analysis of all these qualitative data will be led using an iterative approach in a team and will involve the subsequent reading of the verbatim records, the coding, and the thematic analysis in connection with the framework, followed by validation by the participants and the monitoring committee. One professional and one research assistant will code the interviews according to a mixed strategy that is both deductive (conceptual framework) and inductive (emergence). A summary list of initial codes based on our conceptual framework will serve as an *a priori* coding grid. It will be modified and enriched over the course of the coding and preliminary analyses. The coding will be monitored using a double coding technique carried out by research professional and the main researchers (LM, GCT and NT). Parallel independent coding will be done for the first five interviews, followed by a comparison of results. This process will be repeated until a consensus list of initial codes has been made and once a level of intercoder reliability of over 90% has been reached (52). Data from the semi-structured interviews and the observations will be analyzed thematically. The analytical phases will be interspersed with research team meetings and will involve the input of the monitoring committee. The analyses will be conducted using the software QDA Miner 6 (53).

#### Survey and database analysis

*A priori*, our dependent variables are the effects of MCSP, meaning respect for user needs, user pathways and the performance of HCOs (see fig. 2 and table 2). The independent and intermediary variables are the characteristics of MCSP, for example: access mechanisms, interventions for health promotion/evaluation/research/guidance. These variables will be validated by conducting interviews with participants.

We plan to use reliability scores for the implementation of MCSP, ANOVA tests to compare the contexts for implementation, and multiple and logistical linear regressions to model the effects according to the tracer cases (the monitored pathways in each of the HCO (N = 8) and the contexts for implementation). The variables adjusted following the interviews will be used to carry out the analysis. The questionnaire will be sent online via the LimeSurvey^1^ platform. The software that will be used for the treatment and analysis of the data collected by questionnaire and the data collected through the clinical-administrative databases is SPSS 26.0 (54).

Lastly, to go beyond a “universal” definition of management and organizational theories, differentiated analysis according to gender will be included both in terms of the description of the implementation of MCSP (Objectives 1 and 2) but also in terms of the effects and impact (Objectives 3 and 4) as well as the comparison of the contexts for implementation (Objective 5). Differentiated analysis according to gender provides a more representative vision of the population (here: users, community actors, clinicians, professional, decision-makers and managers), considering the differentiated realities and needs of women and men (55, 56). For example, the questionnaire will be formulated in a way that allows differentiation between responses according to gender (accompanied by other variables such as leadership, management and collaboration models, the composition of operational, tactical, and strategic committees, etc.). Similarly, during the qualitative analysis, the corpus of texts will be coded and worked on while taking into consideration this major variable, including in the field of management in HCOs.

## Ethical considerations and declarations

The research protocol was reviewed and approved by Ethics Committee of the institution of health and social services on 28th January 2021 (2021-Projet #MP-31-2021-3799).

Consent for publication: As part of the written informed consent, all participants will be asked to give permission for information they provide in interviews to be published in an anonymized form.

### Status and timeline of the study

Step 1 of the study is under way. Surveys (organizational, partners and users) were developed by the research team on Lime Survey software and tested with a panel of 5 key respondents in December 2022. The time 1 for the survey will be scheduled at the end of February 2023. Next one will be in December 2023 (Time 2), September 2024 (Time 3) and February 2025 (Time 4).

In parallel, we finish to organise the qualitative data collect with 16 individuals from each territory (4) and 6 to 9 individuals from provincial administration and other Canadian provinces. All these persons will be recruited and interviewed between October 2022 and Mars 2025.

Since 2022, we have obtained permission to use the anonymized clinical-administrative database of HCOs in the study. In partnership, we will use these databases and support the implementation of CMHP in each territory through tailored feedback workshops.

## Discussion

### What contribution can we make to MCSP?

Although several studies have demonstrated the impact of the context at the local level (57, 58) and at the national level (59, 60), it is necessary to bring these two research areas together to gain a detailed understanding of the co-evolution between MCSP, the policies and the local actors in terms of a multilevel, adaptive governance system (61).

The call for more collaborative and participatory research, notably involving users and community partners (e.g., community organizations, municipalities, schools) from a systemic perspective in healthcare system, forms part of our theoretical considerations surrounding MCSP. This is particularly relevant (but not only) to the development of knowledge about the adaptation of the MCSP within HCOs in the turbulent environment we are experiencing with the COVID-19 pandemic. The pandemic has had several impacts on the health system (e.g., on resources, service delivery, management, and governance).

In this context, this study could help to increase knowledge of MCSP implementation in different HCOs and their networks, as well as transferability at the provincial level, an aspect that has not been addressed in previous studies. Work on scaling up innovation is gaining momentum (62-65). However, the implementation of governance innovations such as GTSS appears to be promising but less explored. This is exactly the challenge we wish to take up with our partners in order to find the adapted and adaptive formula for each of the territories and trajectories targeted by this study. This study could help to demonstrate how, through MCSP implementation, the health care system can enable health and social service organizations to better adapt to the needs of the population and may has contributed to the ability of HCOs to adapt to the COVID-19 pandemic;

Finally, this study could help to generate practical knowledge by creating theory-based implementation suggestions through the Complex Adaptive System developed together with key stakeholders like HCOs, users, community organisation, municipalities, education sector.

In brief, the implementation of MCSP and its evaluation can be challenging (time, human resources, collaboration), but also generate unprecedented and as yet unknown outcomes from a theoretical (reconciling the local and the political within coherent multi-level adaptive governance), practical (managing uncertainty by collaborating intra- and inter-organizational to meet the needs of populations adaptively), and finally political (building a robust and adaptive health system at the same time is a dream for many governments and societies) perspective.

## Data Availability

No datasets were generated or analysed during the current study. All relevant data from this study will be made available upon study completion.

## Abbreviation

ALC: Alternate level of care
CAS: Complex Adaptive System
CFIR: Consolidated Framework for Implementation Research
ENAP: National School of Public Administration
EPA: European Pathway Association
HCOs: Healthcare Organization(s)
i-CLSC: Data base used in part of HCO (Local Community Service Centers)
IUPLSSS: University Institute on primary health care and social services
MCSP: Management through Care and Services Pathways
MSSS: Ministère de la Santé et des Services sociaux (Ministry of Heath and Social Services)
PAM: Performance Analysis Model
RTS: Réseaux territoriaux et locaux de santé (Territorial and local service networks)

## Author contributions

**Conceptualization**: Lara Maillet, Georges Charles Thiebaut

**Methodology**: Lara Maillet, Georges Charles Thiebaut, Sabina Abou-Malham and Nassera Touati

**Project administration:** Lara Maillet and Anna Goudet

**Writing – original draft**: Lara Maillet, Anna Goudet

**Writing – review & editing:** Lara Maillet, Georges Charles Thiebaut,, Anna Goudet, Nassera Touati, Pernelle Smits, Arnaud Duhoux, Mylaine Breton, Sabina Abou-Malham, Yves Couturier, Frédéric Gilbert, Jean-Sébastien Marchand, Jean-Louis Denis

## Acknowledgment

We would like to thank Elizabeth Hutchings for her assistance in providing translation and language editing services. We also would like to thank Alain Rondeau, retired professor and Paul Lamarche (passed away in February 2021) for their precious advice and all non-authors co-investigators for their implications for this project. We would like to thank Aurelle Jouego, Geneviève Champagne and Gisèle Ntanda for your precious help and support for revisions after useful commentaries from the two reviewers and editor of the journal. Thanks to all!

## Footnotes

1 https://www.limesurvey.org/

